# Inactivating PLEKHA6 Mutations Cause Idiopathic Hypogonadotropic Hypogonadism Through Impaired Kisspeptin Secretion

**DOI:** 10.64898/2026.04.10.26349358

**Authors:** A. Kemal Topaloglu, Lacey Plummer, Chien-Wen Su, Leman Damla Kotan, Gamze Çelmeli, Enver Simsek, Yuhang Zhao, Maria Stamou, Ahmet Anik, Esra Döğer, Selda Ayça Altıncık, Eda Mengen, A. Filiz Koc, Gamze Akkus, Ravikumar Balasubramanian, Ihsan Turan, Stephanie B. Seminara, Bilgin Yuksel

## Abstract

**Purpose:** Idiopathic hypogonadotropic hypogonadism (IHH) is characterized by impaired reproductive maturation, and approximately half of all cases lack an identified genetic cause. We investigated the genetic basis of IHH in two large cohorts to identify novel disease-causing genes.

**Methods:** We analyzed exome and genome sequencing data from 1,822 patients with IHH from two independent cohorts. Rare variants were filtered using pedigree-informed inheritance models. PLEKHA6 expression in the postmortem human hypothalamus were tested at the mRNA and protein level. Functional studies assessed kisspeptin secretion in cell-based assays.

**Results:** We identified 18 distinct *PLEKHA6* variants in 24 patients from 20 unrelated families (1.3% of cohort). Variants segregated with disease under autosomal recessive and autosomal dominant (with variable penetrance) inheritance patterns. PLEKHA6 was robustly expressed in the hypothalamus and showed clear colocalization with neurokinin B, which served as the marker for the GnRH pulse generator. Functional studies demonstrated that patient variants significantly impaired kisspeptin secretion.

**Conclusion:** PLEKHA6 is a novel IHH gene and the first reported regulator of kisspeptin secretion from the kisspeptin-neurokinin B-dynorphin (KNDy) neurons, which have recently been established as the GnRH pulse generator. These findings establish impaired kisspeptin release as a new disease mechanism in IHH and highlight the critical role of neuropeptide trafficking in reproductive function.

## INTRODUCTION

Idiopathic hypogonadotropic hypogonadism (IHH) is a rare genetic disorder characterized by absent or incomplete pubertal development due to deficient gonadotropin-releasing hormone (GnRH) signaling. The condition imposes significant psychosocial burden on affected individuals and is often clinically confused with constitutional delay of growth and puberty. This leads to prolonged diagnostic uncertainty and delayed treatment, often resulting in insufficient secondary sexual characteristics and impaired fertility ^1^. Despite the identification of approximately 60 causal genes, about half of IHH cases remain genetically unexplained, highlighting the need to discover novel disease genes.

We previously identified mutations in genes encoding KNDy neuropeptides—KISS1 (kisspeptin) and TAC3 (neurokinin B)—and their respective receptors KISS1R and TACR3 as causes of IHH ^2-5^. These discoveries were essential in the recent establishment of the kisspeptin-neurokinin B-dynorphin (KNDy) neurons in the arcuate nucleus (ARC) of the hypothalamus as the GnRH pulse generator ^6-8^. KNDy neurons form an oscillating network that rhythmically releases kisspeptin to stimulate pulsatile GnRH secretion from GnRH neurons, thereby driving gonadotropin release from the pituitary ^9-11^.

While the stimulation of KNDy neurons and subsequent synthesis of kisspeptin is relatively well characterized, the secretory mechanisms spanning from the Golgi to plasma membrane responsible for eventual release of kisspeptin remain largely unexplored. The remarkable rhythmic synchrony of kisspeptin release from KNDy neurons suggests a highly specialized secretory apparatus that may serve as an important control node for reproductive function ^12; 13^. Kisspeptin, like other neuropeptides, is packaged into large dense-core vesicles and released through regulated exocytosis—a process requiring coordinated vesicular trafficking from the trans-Golgi network to the plasma membrane ^14; 15^

Here we report the identification of inactivating variants in *PLEKHA6*, encoding a pleckstrin homology (PH) domain-containing protein involved in neuropeptide trafficking, in patients with IHH. Our clinical, genetic, and functional data indicate *PLEKHA6* as a novel IHH gene and reveal impaired kisspeptin secretion as a previously unrecognized mechanism underlying reproductive failure.

## MATERIALS AND METHODS

### Patient Cohorts and Variant Identification

We analyzed whole-exome and genome sequencing data from 1,822 patients with IHH, representing two large cohorts (513 patients from the Mass General Brigham for Children Pediatric Endocrinology cohort and 1,309 patients from the Harvard Reproductive Endocrine Unit cohort). Human experimental protocols were approved by the ethics committee of the Cukurova University Faculty of Medicine and the institutional review board of Mass General Brigham for the Pediatric Endocrinology cohort and the institutional review board of Mass General Brigham for the Harvard Reproductive Endocrine Unit cohort. All individuals and/or their legal guardians provided written informed consent. IHH was diagnosed based on absent or incomplete pubertal development, low sex steroid concentrations, and low or inappropriately normal gonadotropin levels in the absence of anatomical or functional abnormalities of the hypothalamic-pituitary region. Rare variants (minor allele frequency <0.001 in gnomAD or the origin population) were filtered using a web-based analysis tool ^16^. Filtered variants were evaluated by a group of established *in silico* analysis tools: CADD, GERP, PolyPhen-2, and SIFT.

### Clinical Phenotyping

Detailed clinical and hormonal evaluations were performed for all probands and available family members. Assessments included physical examination, and hormonal measurements (LH, FSH, testosterone, estradiol).

### RT-PCR and Immunofluorescence

Total RNA was isolated from fresh-frozen human hypothalamic tissue samples using the RNeasy Mini Kit (Qiagen) according to the manufacturer’s instructions. RNA concentration and purity were assessed spectrophotometrically by Nanodrop. First-strand cDNA was synthesized from total RNA using the RevertAid First Strand cDNA Synthesis Kit (Thermo Fisher Scientific, K1621) following the manufacturer’s protocol. PCR amplification was performed using Platinum™ II Hot-Start PCR Master Mix (Thermo Fisher Scientific, 14001013) with gene-specific primers.

PCR products were resolved by agarose gel electrophoresis and visualized using a nucleic acid stain. For immunofluorescence colocalization experiments fresh-frozen human hypothalamic tissue samples from the NIH NeuroBioBank were sectioned at 10 μm thickness. Sections were fixed, permeabilized, blocked, and incubated with primary antibodies against PLEKHA6 (Rat RtSzr127, a gift from Dr. Sandra Citi)^17^ and neurokinin B (Rabbit NB300-201SS, Novus). Following subsequent incubation with Alexa Fluor 488-conjugated anti-rat (A21208, Invitrogen) and Alexa Fluor 568-conjugated anti-rabbit (A11011, Invitrogen) antibodies sections were mounted and imaged using Olympus Fluoview FV3000 Laser Scanning Confocal w/IX83 P2ZF microscopy. Colocalization analysis was performed using the Coloc 2 plugin in Fiji ^18^. Pearson’s correlation coefficient and Manders’ split coefficients (M1 and M2)^19^ were calculated following Costes’ automatic threshold regression ^20^. Statistical significance of colocalization was assessed using Costes’ randomization test with 200 iterations.

### Cell Culture and Transfection

Human embryonic kidney (HEK293T) cells were cultured in DMEM supplemented with 10% fetal bovine serum and 1% penicillin-streptomycin at 37°C with 5% CO_2_. Cells were transfected using Lipofectamine 2000 according to manufacturer’s instructions.

### Western Blotting

Protein from HEK293T cell lysates expression was assessed by Western blotting. Proteins were separated by SDS-PAGE, transferred to PVDF membranes, blocked, and incubated with primary (rabbit anti-DDK antibody, 20543-1-AP Proteintech and mouse anti-β-actin MA1-140, Thermo Fisher) and fluorescent secondary (Goat Anti-Rabbit 12004158 and Goat Anti-mouse 12005866, Bio-Rad) antibodies. Signals were detected using fluorescence imaging.

### Kisspeptin Secretion Assays

HEK293T cells were transfected with constructs encoding pre-pro-KISS1 (a gift from Dr. Lomniczi, OHSU) along with empty vector (EV), wild-type PLEKHA6 (RC218402 Origene) or engineered mutant versions (Genescript). At 48 hours post-transfection, media were collected, clarified by centrifugation, and stored at -80°C. Kisspeptin levels were measured using a commercial ELISA kit (ab288589 Abcam) according to manufacturer’s instructions. Results were normalized to total protein content measured by BCA assay.

### Statistical Analysis

Data are presented as mean ± standard error of the mean (SEM). Statistical comparisons were performed using SPSS. P values <0.05 were considered statistically significant. All experiments were performed in at least three independent biological replicates.

## RESULTS

### Identification of PLEKHA6 Variants in IHH Patients

Through systematic analysis of whole-exome and genome sequencing data from 1,822 IHH patients, we identified 18 distinct *PLEKHA6* variants in 24 patients from 20 independent families (Table 1). These missense variants distributed across the PLEKHA6 gene, affecting multiple functional domains including the pleckstrin homology (PH) domain, proline-rich region, coiled-coil domain, and C-terminal region (Fig 1). All variants were rare (minor allele frequency <0.001 in gnomAD). *PLEKHA6* variants accounted for approximately 1.3% of cases in our cohort, a frequency comparable to most of the 60 previously known IHH genes. Notably, mutations in KISS1 and TAC3, which encode two of the most critical KNDy neuropeptides, each occur in fewer than 2% of IHH cases^21^.

**Table 1.** The molecular genetic characteristics of the PLEKHA6 variants. Abbreviations: Het, heterozygous; Hom, homozygous; MAF, minor allele frequency; GnomAD, The Genome Aggregation Consortium; CADD, Combined Annotation Dependent Depletion; GERP, Genomic Evolutionary Rate Profiling; ACMG/AMP, the American College of Medical Genetics and Genomics and the Association for Molecular Pathology; VUS, variant uncertain significance; PM, pathogenic moderate; PP, pathogenic supporting; BP, benign supporting; PolyPhen-2, Polymorphism Phenotyping v2; SIFT, Sorting Intolerant From Tolerant; D, deleterious; B, benign; T, tolerated; N/A, not available; Variants are described according to the RefSeq numbers: *PLEKHA6*, NM_014935; *PLEKHA5*, NM_001256470; *DMXL2*, NM_001378457; *PROK2*, NM_001126128; *GNRH1*, NM_001083111; *ANOS1*, NM_000216; and *TACR3*, NM_001059.

| Family/Individual no | Variant at cDNA level | Variant at protein level | Zygosity | GnomAD MAF | CADD score | GERP | PP2 | SIFT | ACMG/AMP | Additional known/candidate IHH gene variant |
| --- | --- | --- | --- | --- | --- | --- | --- | --- | --- | --- |
| A-1 | c.836G>A | p.Arg279Gln | Hom | 0,00004 | 28,2 | 5,5 | D | D | VUS: PM1 |  |
| A-2 |  |  |  |  |  |  |  |  |  |  |
| A-3 |  |  |  |  |  |  |  |  |  |  |
| B-1 | c.628G>A | p.Glu210Lys | Het | 0,00001 | 25,8 | 5,5 | D | T | VUS: PM1, PM2 |  |
| B-2 |  |  |  |  |  |  |  |  |  |  |
| C-1 | c.1578G>T | p.Asn526Lys | Het | 0,00089 | 20,8 | 2,6 | D | D | VUS: PM1 |  |
| C-2 |  |  |  |  |  |  |  |  |  |  |
| D-1 | c.2546C>T | p.Pro849Leu | Het | 0,00001 | 24,0 | 5,2 | D | T | VUS: PM1, PM2 |  |
| E-1 | c.3074C>T | p.Pro1025Leu | Het | 0,00002 | 19,6 | 1,3 | B | T | VUS: PM1, BP4 |  |
| F-1 | c.2642G>A | p.Arg881Gln | Het | 0,00006 | 32,0 | 5,5 | D | D | VUS: PM1 | *PLEKHA5* p.Thr884Ile |
| G-1 | c.2507T>C | p.Met836Thr | Het | 0,00000 | 21,0 | 4,1 | B | D | VUS: PM1, PM2 | *DMXL2* p.Phe2629Cys |
| H-1 | c.2716G>C | p.Glu906Gln | Het | 0,00005 | 24,4 | 5,4 | D | T | VUS: PM1 |  |
| I-1 | c.3123C>G | p.Asp1041Glu | Het | 0 | 25,9 | 4,5 | D | D | VUS: PM1, PM2 |  |
| K-1 | c.2786G>A | p.Arg929Gln | Het | 0 | 33,0 | 5,3 | D | D | VUS: PM1, PM2 | *PROK2* p.Arg73Cys |
| L-1 | c.2417C>T | p.Pro806Leu | Het | 0 | 25,8 | 5,2 | D | D | VUS: PM1, PM2 | *GNRH1*, p.Arg262Gln |
| M-1 | c.2167A>G | p.Asn723Asp | Het | 0 | 18,7 | 4,5 | B | T | VUS: PM1, PM2 | *ANOS1* p.Gln100Lys |
| N-1 | c.1801G>A | p.Val601Met | Het | 0,00001 | 24,8 | 4,8 | D | T | VUS: PM1, PP3 |  |
| O-1 | c.1318G>A | p.Ala440Thr | Het | 0,00003 | 16,3 | 2,5 | B | T | VUS: PM1 | *TACR3* p.Arg230Cys |
| P-1 | c.848C>T | p.Thr283Ile | Het | 0 | 23,4 | 4,3 | D | T | VUS: PM1 |  |
| Q-1 | c.836G>A | p.Arg279Gln | Het | 0,00004 | 28,2 | 5,5 | D | D | VUS: PM1 |  |
| R-1 | c.581C>A | p.Pro194His | Het | 0 | 14,5 | 3,6 | B | T | VUS: PM1, PM2 |  |
| S-1 | c.565C>G | p.His189Asp | Het | 0,00009 | 19,9 | 5,6 | B | T | VUS: PM1 |  |
| T-1 | c.247G>A | p.Val83Ile | Het | 0,00008 | 26,2 | 5,5 | D | D | VUS: PM1 |  |
| U-1 | c.247G>A | p.Val83Ile | Het | 0,00008 | 26,2 | 5,5 | D | D | VUS: PM1 |  |

**Figure 1.**
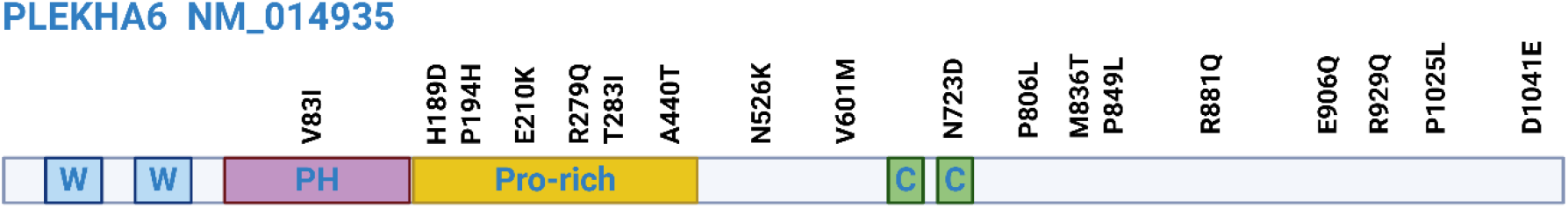
PLEKHA6 variants in patients with IHH. Distribution of 18 different PLEKHA6 variants across the gene is shown.

### Segregation Analysis

One multiplex family demonstrated an autosomal recessive variant inheritance pattern. Another multiplex family provided *de novo* evidence likely due to gonadal mosaicism. The inheritance in the other families is consistent with autosomal dominant pattern with variable penetrance and expressivity, a phenomenon commonly observed in IHH families ^22; 23^.

All probands presented with absent or incomplete pubertal development. Males exhibited absent or severely impaired testicular enlargement, insufficient virilization, and prepubertal testosterone levels. Females showed absent breast development, primary amenorrhea, and low estradiol levels. Gonadotropin levels (LH and FSH) were low or inappropriately normal for the degree of sex steroid deficiency, consistent with central hypogonadism. In one patient we were able to assess the GnRH response to extended daily GnRH injections for seven days. He showed normal gonadotropin response in GnRH stimulation test immediately after the treatment period suggesting the hormonal deficit for IHH is located in the hypothalamus, not the pituitary ^24^.

The molecular genetic characteristics of the *PLEKHA6* variants are shown in Table 1. All variants were classified as variant of uncertain significance (VUS), by ACMG/AMP classification ^25^. However, Polyphen-2 ^26^ and SIFT ^27^, two well-validated in silico prediction programs, indicated the great majority of these variants to be harmful. The variants were either not seen in the largest reference population database (gnomAD) or occurred at an extremely rare minor allele frequency of <0.0005. Some of the variants were previously reported in ClinVar as VUS. No other potentially harmful variants in the known IHH-associated genes other than those listed in Table 1 was found. Notably, the proband in one family had an additional variant in *PLEKHA5*, which is the closest paralog of *PLEKHA6*. Previously, in another study a *PLEKHA5* variant was preliminarily reported to cause IHH in one patient ^28^. In the rest of these pedigrees there could be additional variants in yet unknown IHH-associated genes as only half of the IHH cases are explained by variants in the known IHH genes^29^.

### PLEKHA6 Expression in the Hypothalamus

Single-cell RNA sequencing studies in mice indicate that Plekha6 is highly expressed in the hypothalamus, enriched in embryonic ARC clusters ^30^, and present in the ARC Kiss1-high subcluster across pubertal developmental stages ^31^. These expression patterns are consistent with a functional role in the GnRH pulse generator. To investigate whether PLEKHA6 is expressed in relevant neuronal populations in humans, we performed Rt-PCR and immunofluorescence studies on fresh-frozen human hypothalamic tissues. At the mRNA level PLEKHA6 is expressed along with known KNDy neuron genes including, KISS1, TAC3, TACR3, and PDYN (Fig 2 Upper panel). Confocal microscopy analysis of hypothalamic sections from three young adult males, revealed strong colocalization between PLEKHA6 and NKB. Quantitative analysis of 15 images (5 images per subject) using Coloc 2 in Fiji demonstrated a Pearson’s correlation coefficient of 0.738 ± 0.109, with Manders’ split coefficients of M1 = 0.702 ± 0.163 and tM2 = 0.790 ± 0.134, indicating that approximately 73% of PLEKHA6 signal overlapped with NKB and 79% of NKB signal overlapped with PLEKHA6. Costes’ randomization test confirmed that observed colocalization was statistically non-random (p > 0.95). Hence PLEKHA6 protein was robustly expressed in the hypothalamus and showed clear colocalization with NKB, which served as a proxy marker for KNDy neurons due to lack of validated anti-human kisspeptin antibodies (Fig 2 Lower panel). This expression and colocalization pattern support the hypothesis that PLEKHA6 functions in kisspeptin-secreting neurons.

**Figure 2.**
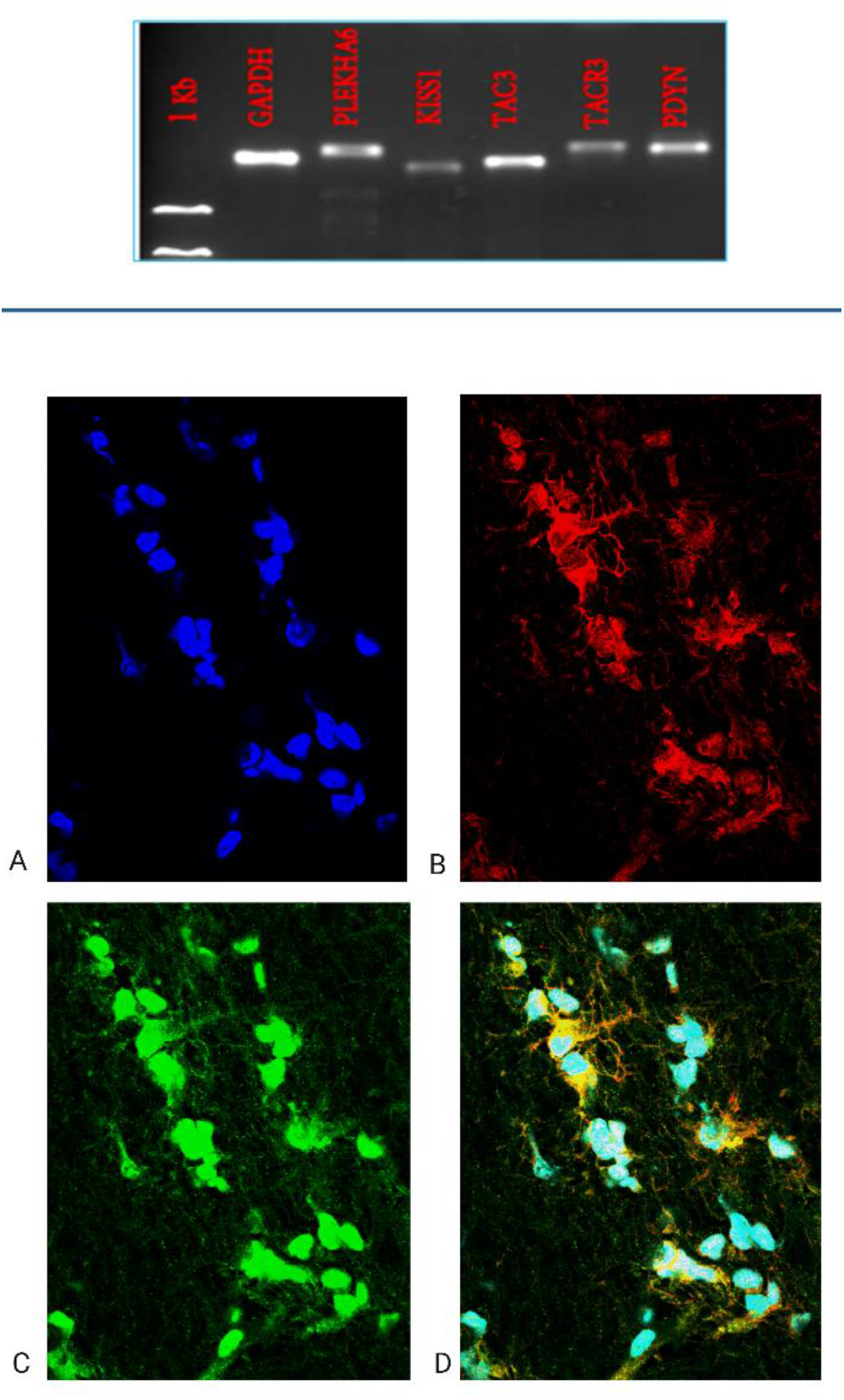
PLEKHA6 expression in human hypothalamic KNDy neurons. Upper panel: RT-PCR of fresh-frozen human hypothalamic tissue demonstrates expression of PLEKHA6 alongside established KN Dy neuronal markers, including kisspeptin (KISS1), neurokinin B (TAC3), neurokinin B receptor (TACR3), and prodynorphin (PDYN), confirming transcriptional expression of PLEKHA6 within the KN Dy neuronal population. Lower panel: Representative confocal immunofluorescence images of fresh-frozen postmortem human hypothalamic tissue acquired using an Olympus FV3000 confocal microscope (60× objective). (A) DAPI nuclear counterstain, (B) Neurokinin B, (C) PLEKHA6, and (D) Merged image. The merged image demonstrates colocalization of PLEKHA6 and Neurokinin B signals. Images are representative of n = 3 subjects.

### Patient Variants Impair Kisspeptin Secretion

To assess whether patient variants affected the protein expression, we performed western blotting in five selected variants across the gene. The results showed comparable expression patterns to that of the WT (Fig 3A). Then to test the functional consequences of PLEKHA6 variants, we performed kisspeptin secretion assays in HEK293T cells co-transfected with pre-pro-kisspeptin and PLEKHA6 constructs. Representative patient variants from Family A (p.R279Q) and Family B (p.E210K) were tested. Kisspeptin concentrations were significantly lower from the patient variants in comparison to WT (Fig 3B). These results indicate that the patient variants are loss-of-function alleles and impair kisspeptin release from KNDy neurons, disrupting a critical upstream neuropeptide signal to stimulate GnRH neurons.

**Figure 3.**
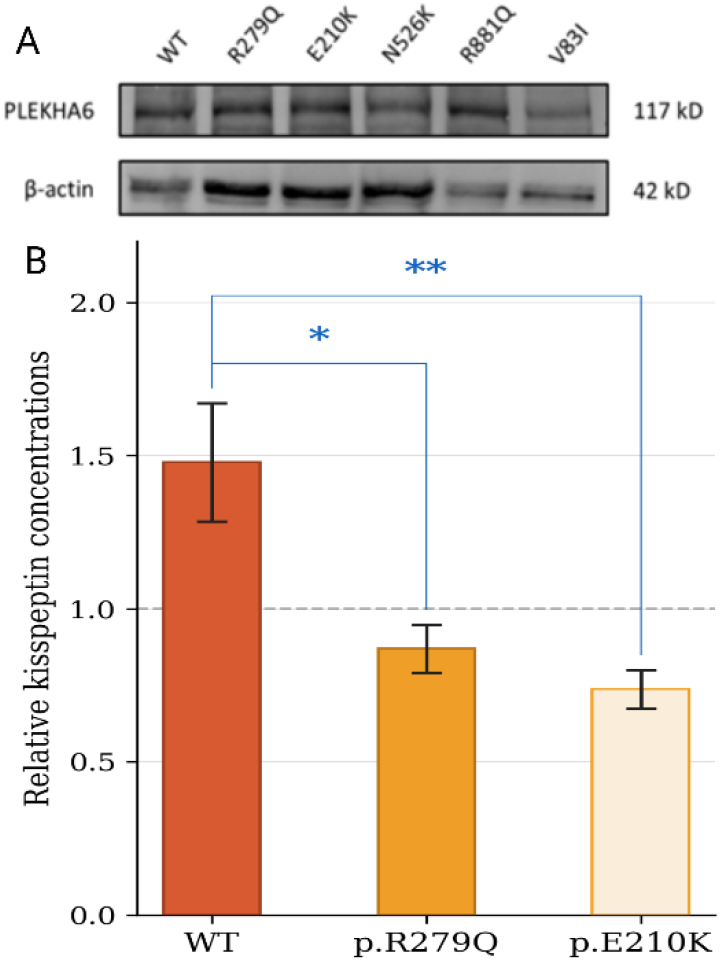
Patient PLEKHA6 variants impair kisspeptin secretion. (A) Western blot analysis of five selected variants across the gene transfected into HEK293T cells did not show reduced expression. (B) Kisspeptin levels were measured by ELISA in media collected from HEK293T cells co-expressing pre-pro-kisspeptin with wild-type PLEKHA6 (WT), the Family A variant (p.Arg279Gln) or the Family B variant (p.Glu210Lys). Kisspeptin levels were normalized to total protein content and expressed relative to EV (dashed line). Statistical analyses were performed using the Kruskal–Wallis test, followed by Dunn’s post hoc test with Bonferroni correction (SPSS). Data represent n = 19 independent biological replicates. Error bars indicate SEM. *p < 0.05 vs. WT; **p <0.01 vs. WT.

## DISCUSSION

We report inactivating variants in *PLEKHA6* as a novel genetic cause of IHH, identifying impaired kisspeptin secretion as a previously unrecognized disease mechanism. Our findings reveal that PLEKHA6 plays an essential role in mediating kisspeptin release from KNDy neurons, the neuronal population that serves as the GnRH pulse generator. The 60 previously known IHH genes primarily affect GnRH neuron development and migration (e.g., ANOS1, CHD7, FGF8), neuropeptide synthesis (KISS1, TAC3), or receptor signaling (KISS1R, TACR3). Thus, PLEKHA6 represents the first reported regulator of the secretory step— specifically, the trafficking and exocytosis of kisspeptin-containing vesicles. Pulsatile kisspeptin secretion is a remarkably coordinated physiological event, requiring the simultaneous and synchronized release of kisspeptin across a bilaterally distributed network of KNDy neurons in the ARC of the hypothalamus. This extraordinary degree of synchrony demands highly specialized secretory machinery capable of meeting both the high-capacity and temporally coordinated kisspeptin release. Our findings demonstrate that PLEKHA6 is a critically important trafficking protein in this distinct and genetically vulnerable node subject to disruption. Recent work by Citi and colleagues demonstrated that PLEKHA6 regulates recycling of ATP7A-containing vesicles to maintain copper homeostasis, coordinating through cooperative interactions among its PH, proline-rich, and coiled-coil domains. ^17; 32^. Our findings extend this model to neuroendocrine secretion, establishing vesicle trafficking in KNDy neurons as a new mechanistic category in the genetics of IHH.

The identification of PLEKHA6 as an IHH gene has several important clinical implications. First, it expands the genetic testing panel for patients with unexplained pubertal delay and infertility. Given that PLEKHA6 accounts for 1.3% of cases in our cohort, screening this gene should be incorporated into clinical diagnostic panels. Second, the variable penetrance and oligogenic architecture observed in our families underscore the complexity of IHH genetics and the need for comprehensive genetic evaluation. Finally, the identification of PLEKHA6 as a regulator of kisspeptin secretion opens new avenues for manipulation and intervention.

Our study has limitations. First, while our functional studies clearly demonstrate impaired kisspeptin secretion in cell-based assays, these experiments were performed in HEK293T cells which may not fully recapitulate the specialized secretory machinery of KNDy neurons. Second, not all patients carrying PLEKHA6 variants manifested IHH, consistent with variable penetrance. This suggests that additional variants in yet unidentified IHH genes, epigenetic factors, or environmental influences may modulate disease expression. Finally, we do not present data from animal models —currently lacking—which will be essential for studying the in vivo consequences of PLEKHA6 deficiency.

## Conclusions

We have identified PLEKHA6 as a novel IHH gene and established impaired kisspeptin secretion as a previously unrecognized disease mechanism. Our findings reveal that regulated neuropeptide secretion represents a critical, genetically vulnerable control node in the reproductive axis. These discoveries expand the genetic landscape of IHH, reveal new biology of reproductive control, and open opportunities for modulating kisspeptin output.

## Data Availability

Anonymized genetic variant data will be deposited in ClinVar. Additional data supporting the findings of this study are available from the corresponding author upon reasonable request.

https://www.ncbi.nlm.nih.gov/clinvar/

## ACKNOWLEDGMENTS

We thank the patients and families who participated in this study. We thank Dr. Sandra Citi of University of Geneva for invaluable discussions. We thank the NIH NeuroBioBank for providing human hypothalamic tissue samples. S.B.S. is supported by grants P50HD104224 (National Center for Translational Research in Reproduction and Infertility) and R37HD043341 from the Eunice Kennedy Shriver National Institute of Child Health and Development and grant R01FD007843 from the FDA. She is also supported by the Gates Foundation.

